# Emergence and ongoing outbreak of ST80 vancomycin-resistant *Enterococcus faecium* in Guangdong province, China from 2021 to 2023: a multicenter, time-series and genomic epidemiological study

**DOI:** 10.1101/2024.03.20.24304567

**Authors:** Cong Shen, Li Luo, Hongyun Zhou, Yinglun Xiao, Jinxiang Zeng, Liling Zhang, Jieying Pu, Jianming Zeng, Ni Zhang, Yueting Jiang, Lingqing Xu, Dingqiang Chen, Gang Li, Kuihai Wu, Hua Yu, Min Wang, Xuemin Guo, Juan Wang, Bin Huang, Cha Chen

## Abstract

**Background:** The surveillance system revealed that the prevalence of vancomycin-resistant *Enterococcus faecium* (VREfm) has increased. We aim to investigate the epidemiological and genomic characteristics of VREfm in China.

**Methods:** We collected 20747 non-redundant *E. faecium* isolates from inpatients across 19 hospitals in six provinces between Jan 2018 and June 2023. VREfm was confirmed by antimicrobial susceptibility testing. The prevalence was analyzed using changepoint package in R. Genomic characteristics were explored by whole-genome sequencing and bioinformatic analysis.

**Results:** 5.59% (1159/20747) of *E. faecium* isolates were resistant to vancomycin. The prevalence of VREfm increased in Guangdong province from 5% before 2021 to 20%-50% in 2023 (p<0.0001), but not in the other five provinces. The two predominant clones before 2021, ST17 and ST78, were substituted by an emerging clone, ST80, from 2021 to 2023 (88.63%, 195/220). All ST80 VREfm from Guangdong formed a single lineage (SC11) and were genetically distant from the ST80 VREfm from other countries, suggesting a regional outbreak. All ST80 VREfm in SC11 carried a new type of plasmid which harbored a *vanA* cassette (*vanRSHAXYZ*) flanked by Tn*1546*/Tn*3* clusters. However, no conjugation-related gene was detected and no transconjugant was obtained in conjugation experiment, indicating that the outbreak of ST80 VREfm could be attributed to clonal transmission.

**Conclusions:** We revealed an ongoing outbreak of ST80 VREfm with a new *vanA*-harboring plasmid in Guangdong, China. This clone has also been identified in other provinces and countries, foreboding a risk of wider spreading shortly. Continuous surveillance is needed to inform public health interventions.

## 1. Introduction

Vancomycin-resistant *Enterococcus faecium* (VREfm) is one of the leading causes of severe healthcare-associated infections, such as urinary tract infections, intra- abdominal infections and bloodstream infections, which result in high mortality rates and significant burdens of disease on human society [1–4]. Over the past decade, VREfm has emerged in the hospital setting in high prevalence, causing outbreaks and severe infections in Europe, America, and Australia [4–7]. Owing to the limited therapeutic alternatives, VREfm were classified as high-priority pathogens among antibiotic-resistant bacteria by the WHO [2].

In China, the prevalence of VREfm has remained low (average <5%) in the past decades, according to the China Antimicrobial Surveillance Network (CHINET) (http://www.carss.cn/). Despite this low prevalence, VREfm infections are associated with increased morbidity, mortality, healthcare costs and duration of hospital stay compared with vancomycin-susceptible (VSE) infections [8]. The routine surveillance of antimicrobial resistance in our hospital indicated that the prevalence of VREfm has increased since January 2020. However, the epidemiological and genomic characteristics of VREfm remain unknown.

Herein, we collected VREfm isolates in 19 hospitals in six provinces in China from January 2018 to June 2023, aiming to reveal the epidemiological and whole-genome sequencing-based characteristics of the VREfm outbreak in Guangdong province.

## 2. Subjects

We conducted a multicenter and epidemiological survey of *E. faecium* from January 2018 to June 2023, in which 19 tertiary hospitals in China participated (Figure 1).

**Figure 1.**
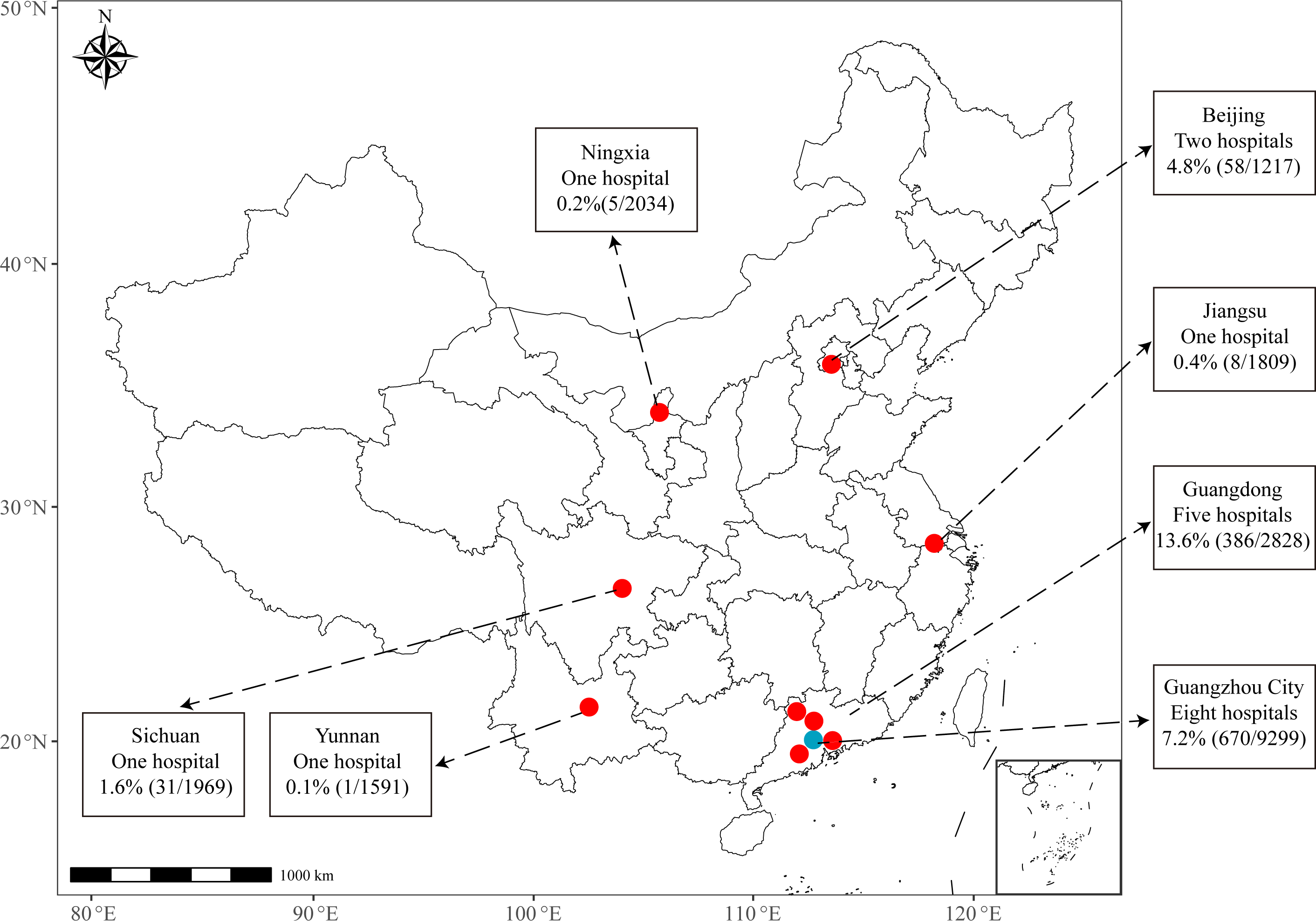
Geographic location of 19 hospitals from six provinces. Data showing in the box are % (n/N). %, prevalence of VREfm. n, Number of VREfm. N, sample size of *E. faecium*.

Among them, eight hospitals are located in Guangzhou city of Guangdong province, five hospitals in other five cities in Guangdong province, and six hospitals in five provinces or municipalities (Yunnan, Jiangsu, Sichuan, Ningxia and Beijing). Subjects were included if clinical samples were evidenced by a positive *E. faecium* culture, and the patients had typical clinical infectious symptoms and abnormal laboratory markers of infections. Patients who exhibited no infectious symptoms were classified as colonisation and were excluded. This study was approved by the ethics committee of Guangdong Provincial Hospital of Chinese Medicine, Guangzhou, China (Ethics number ZE2023-077).

## 3. Materials and methods

### 3.1 Clinical samples and identification of *E. faecium*

Samples from patients with infections with *E. faecium* were collected as part of routine clinical management and/or hospital surveillance. Clinical samples (urine, blood, sputum, wound samples) from patients were plated on Columbia blood agar (CBA) with 5% sheep blood (Luqiao, Beijing, China). Species identification was confirmed by MALDI-TOF MS (Biotyper version 3.2, Bruker Daltonik GmbH, Bremen, Germany).

### 3.2 Antimicrobial susceptibility testing

Vancomycin susceptibility testing was performed using the VITEK 2 (bioMérieuxTM) automated system. Minimum inhibitory concentrations (MICs) for 13 antimicrobials were determined by the broth dilution method.

We extracted the MIC breakpoints used to define resistance and non-susceptibility in *E. faecium* for the thirteen antimicrobials studied, including EUCAST epidemiological cut-offs, EUCAST clinical breakpoints v12.0 and CLSI breakpoints (M100-S33). Non-susceptibility is defined as the category comprising both the intermediate and the resistance categories. EUCAST clinical breakpoints were chosen preferentially. For antibiotics with no EUCAST clinical breakpoints set (i.e. daptomycin, rifampicin, erythromycin, and nitrofurantoin), we used CLSI breakpoints instead (Table 1).

**Table 1.** MIC breakpoints recommended by EUCAST, CLSI, and those used in this study.

| Antibiotics | ECOFF | EUCAST clinical breakpoints |  |  | CLSI breakpoints |  |  | MIC breakpoints chosen in this study |  | Criteria for choosing breakpoint |
| --- | --- | --- | --- | --- | --- | --- | --- | --- | --- | --- |
|  |  | S | I | R | S | I | R | Non-S | R |  |
| Vancomycin | >4 | ≤4 | - | >4 | ≤4 | 8-16 | ≥32 | >4 | >4 | EUCAST CB |
| Teicoplanin | >2 | ≤2 | - | >2 | ≤8 | 16 | ≥32 | >2 | >2 | EUCAST CB |
| Linezolid | >4 | ≤4 | - | >4 | ≤2 | 4 | ≥8 | >4 | >4 | EUCAST CB |
| Tigecycline | >0.25 | ≤0.25 | - | >0.25 | - | - | - | >0.25 | >0.25 | EUCAST CB |
| Daptomycin | >8 | IE | IE | IE | ≤4 | - | ≥8 | >4 | >4 | CLSI CB |
| Rifampicin | ID | - | - | - | ≤1 | 2 | ≥4 | >1 | >2 | CLSI CB |
| Erythromycin | >4 | - | - | - | ≤0.5 | 1-4 | ≥8 | >0.5 | >4 | CLSI CB |
| Ampicillin | >8 | ≤4 | 8 | >8 | ≤8 | - | ≥16 | >8 | >8 | EUCAST CB |
| Ciprofloxacin | >8 | ≤4 | - | >4 | ≤1 | 2 | ≥4 | >4 | >4 | EUCAST CB |
| Levofloxacin | >8 | ≤4 | - | >4 | ≤2 | 4 | ≥8 | >4 | >4 | EUCAST CB |
| Fosfomycin | >128 | - | - | - | ≤64 | 128 | ≥256 | >64 | >128 | CLSI CB |
| Nitrofurantoin | >256 | - | - | - | ≤32 | 64 | ≥128 | >32 | >64 | CLSI CB |
| HGEN | >32 | ≤128 | - | >128 | <500 | - | ≥ 500 | >128 | >128 | EUCAST CB |

EUCAST ECOFF extracted from https://mic.eucast.org/search/; EUCAST clinical breakpoints v13.0 download from https://www.eucast.org/clinical_breakpoints/; CLSI breakpoints extracted from CLSI M100-S33. CB, clinical breakpoint. CLSI, Clinical & Laboratory Standards Institute. ECOFF, epidemiological cut-off; EUCAST, The European Committee on Antimicrobial Susceptibility Testing; IE, insufficient evidence; ID, insufficient data; S, susceptible; I, intermediate; R, resistance; Non-S, non-susceptible; HGEN, high-level gentamycin resistance.

### 3.3 Changepoint analysis of VREfm prevalence

A three-month moving average approach was used to remove noise of monthly VREfm prevalence. The changepoint detection was conducted to identify the significant changes. The function of cpt.meanvar from the changepoint package was used to explore a variety of penalty values and methods, retaining the most consistently identified changepoints with the proposed pruned exact linear time (PELT) algorithm [9].

### 3.4 Whole-genome sequencing and bioinformatic analysis of VREfm isolates

For VREfm collected from Jan 2021 to Jun 2023 in Guangdong, we randomly selected a subset (20%, n=220) of isolates from each month for WGS. For VREfm collected from 2014 to 2020, or from other province, all isolates (n=91) were included for WGS due to its low prevalence. DNA was extracted and sequenced by Illumina Hiseq 4000 platform. Draft genome was assembled using SPAdes v.13.1 [10]. *In silico* multilocus sequence typing (MLST), antimicrobial resistance genes (ARGs), virulence factors (VFs), insertion sequence (IS), and plasmid replicon were established using ABRicate v0.2 [11–13]. Pan-genome analysis was done using Roary v3.11.2, and core genome single-nucleotide polymorphisms (cgSNPs) were extracted using SNP-sites [14, 15]. Phylogeny was constructed by RAxML using cgSNPs [16]. Sequence cluster (SC) was defined using hierBAPS [17]. Five representative isolates that harbored *vanA*-plasmids were sequenced by Illumina PacBio RSII system. The plasmid sequence was circled by Pilon, and plasmid structures were compared by Easyfig [18, 19].

### 3.5 Plasmid conjugation assay

Plasmid conjugation was performed using fusidic acid-resistant *Enterococcus faecium* BM4105 as the recipient. Donor and recipient isolates were cultured overnight and sub-cultured at a 1:100 ratio for 3 hours at 37°C. Then, the donor and recipient were mixed at a 1:9 ratio and incubated stationary for 6 hours at 37_. Transconjugants were selected on BHI agar plates supplemented with fusidic acid (50 mg/L) and vancomycin (8 mg/L), and verified using MALDI-TOF and PCR for *vanA*.

### 3.6 Statistical analyses

Statistical analyses and random selection were performed using R v3.4. Differences in antimicrobial resistance rates were assessed using the Fisher’s exact test. Given that MICs were derived from isolate growth in doubling dilutions of antimicrobials, MIC values were log2-transformed. Differences in distributions of MICs and cgSNPs between groups assessed using Wilcoxon-Mann-Whitney tests. A p-value <0.05 was considered significant.

## 4. Results

### 4.1 The prevalence of VREfm increased in Guangdong Province, China after 2021

Among 19 hospitals from six provinces, 20747 non-redundant *E. faecium* isolates were collected from Jan 2018 to June 2023. Overall, 5.59% (1159/20747, 95% confidence interval [CI]: 5.27%-5.90%) of *E. faecium* isolates were resistant to vancomycin (Figure 1). 54.79% (n=635) of VREfm were collected from urine samples, followed by blood (7.77%, n=90), wound (6.99%, n=81) and sputum (4.49%, n=52). The median age of patients infected with VREfm was 70 years (interquartile range: 57-79).

Time-series analysis showed that the prevalence of VREfm in all hospitals remained low (<5%) before December 2020, except for two hospitals from Beijing were fluctuated up to 20% (Figure 2). Remarkably, the prevalence of VREfm dramatically increased up to 20%-50% after January 2021 (compared with 5% in 2014-2020, p<0.0001) in the hospitals in Guangzhou City and other cities in Guangdong province (Figure 2A and 2B). However, no significant increase was observed in the other five provinces (Figure 2C). According to changepoint analysis, the increasing trend of VREfm prevalence in Guangdong Province started between December 2020 and December 2021 (mostly concentrated on January-June 2021), indicating an ongoing outbreak of VREfm in Guangdong Province during the first half year of 2021 (Figure 2).

**Figure 2.**
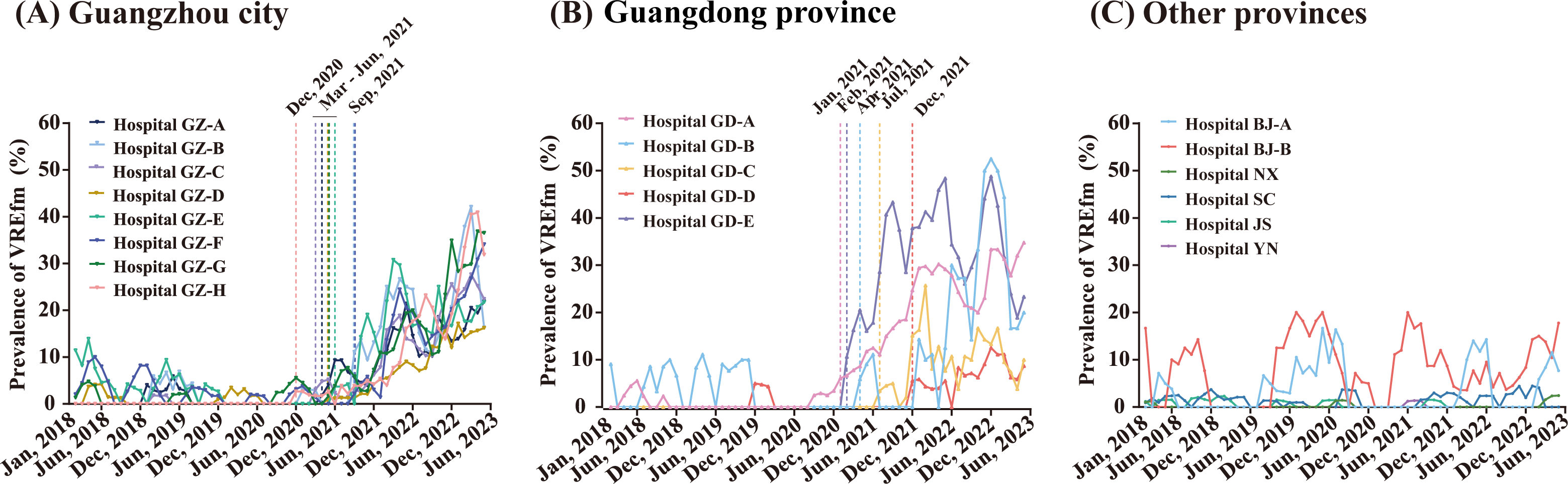
**Time series of monthly VREfm prevalence in Guangzhou city (A), Guangdong province except for Guangzhou city (B) and other five provinces (C), January 2018-June 2023.**The solid line represents the three-month moving average data of VREfm prevalence for each hospital. Vertical dashed lines indicate significant changepoints identified in the changepoint analysis.

### 4.2 MLST shift in dominating clones

To systematically investigate genomic characteristics, we randomly selected 291 VREfm isolates collected from 2018 to 2023 and 20 VREfm isolates from 2014 to 2017 in 19 hospitals for whole-genome sequencing. All isolates (N=311) were typed into 22 sequence types (STs), of which all belong to hospital-associated *E. faecium* clade A1 (Figure 3A and supplement figure 1). The most prevalent STs were ST80, ST78 and ST17, which accounted for 82.99% (n=255) of VREfm. A total of 208 isolates (66.88%) belong to ST80, which have been rarely detected in China [8, 20–22], but commonly detected in Europe, Australia, and America [4–7].

**Figure 3.**
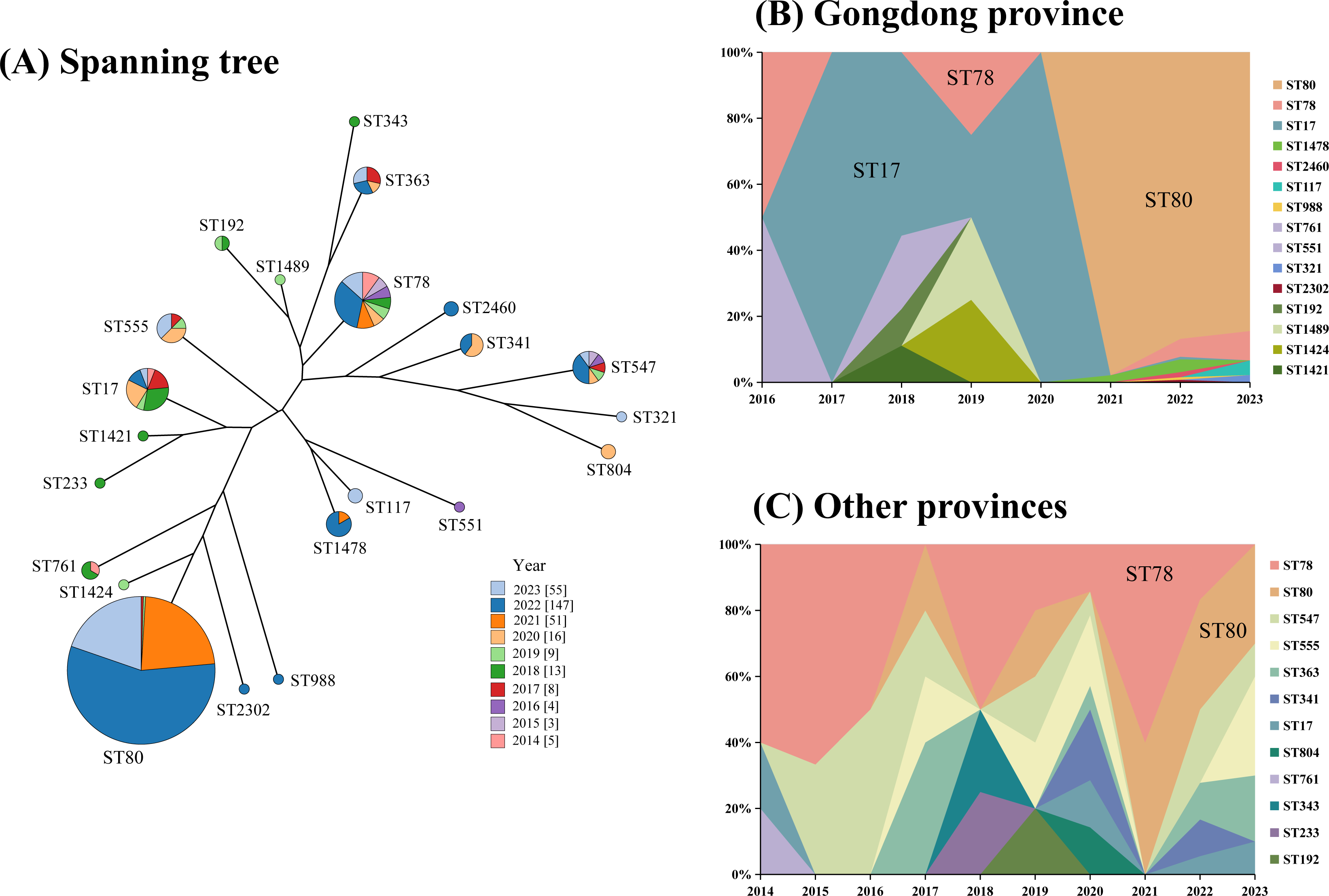
Minimum-spanning tree (A) and dynamic changes (B and C) of MLST distribution from 2014-2023. (A) Each circle corresponds to a unique ST. The number outside the circle indicates ST number. The size of the circle represents the number of isolates belonging to the same ST. The colors inside the circle represent the proportion of the year of isolation. (B) and (C) The x-axis represents the year of isolates. The y-axis represents the proportion of ST.

We discovered that ST17 was predominant in Guangdong Province before December 2020 (55%, 11/20). Remarkably, ST80 appeared in 2021 and became predominant ST from 2021 to 2023 (88.63%, 195/220), suggesting an outbreak of ST80 VREfm in Guangdong Province (Figure 3B). In other provinces, ST78 was the most prevalent ST from 2014 to 2021 (32.56%, 14/43). However, we observed that the predominant ST in 2022 and 2023 has been changed to ST80 (32.14%, 9/28), even though the prevalence of VREfm has not increased (Figure 3C). Our results demonstrated that hospital-associated clade A1 VREfm is consistently endemic in China, while the predominance of ST78 and ST17 could have been replaced by ST80 VREfm, which implied a latent risk that ST80 VREfm could trigger an outbreak in other provinces beyond Guangdong province.

### 4.3 Antimicrobial susceptibility profile of VREfm

Overall, more than 90% of VREfm isolates were non-susceptible to ampicillin (99.0%, n=308), followed by teicoplanin (98.4%, n=306), fosfomycin (98.1%, n=305), ciprofloxacin (97.4%, n=303), and levofloxacin (96.7%, n=301) (Figure 4A). A low non-susceptible rate was observed in linezolid (0.3%, n=1) and tigecycline (9.3%, n=29). Notably, 16.7% (n=52) of VREfm are non-susceptible to daptomycin, which was considered as a significant antimicrobial for treating VREfm. Non-susceptibility rate of rifampin, nitrofurantoin, daptomycin and high concentration gentamycin in ST80 VREfm were significantly higher than other STs (p<0.05, Figure 4B). The distribution of MICs for vancomycin, teicoplanin, daptomycin, rifampin, fosfomycin, nitrofurantoin and high concentration gentamycin among ST80 VREfm were significantly higher than non-ST80 (p<0.05, supplement figure 2). In summary, the antimicrobial resistance rate and MIC of most antimicrobials were higher in ST80 VREfm isolates than non-ST80 STs.

**Figure 4.**
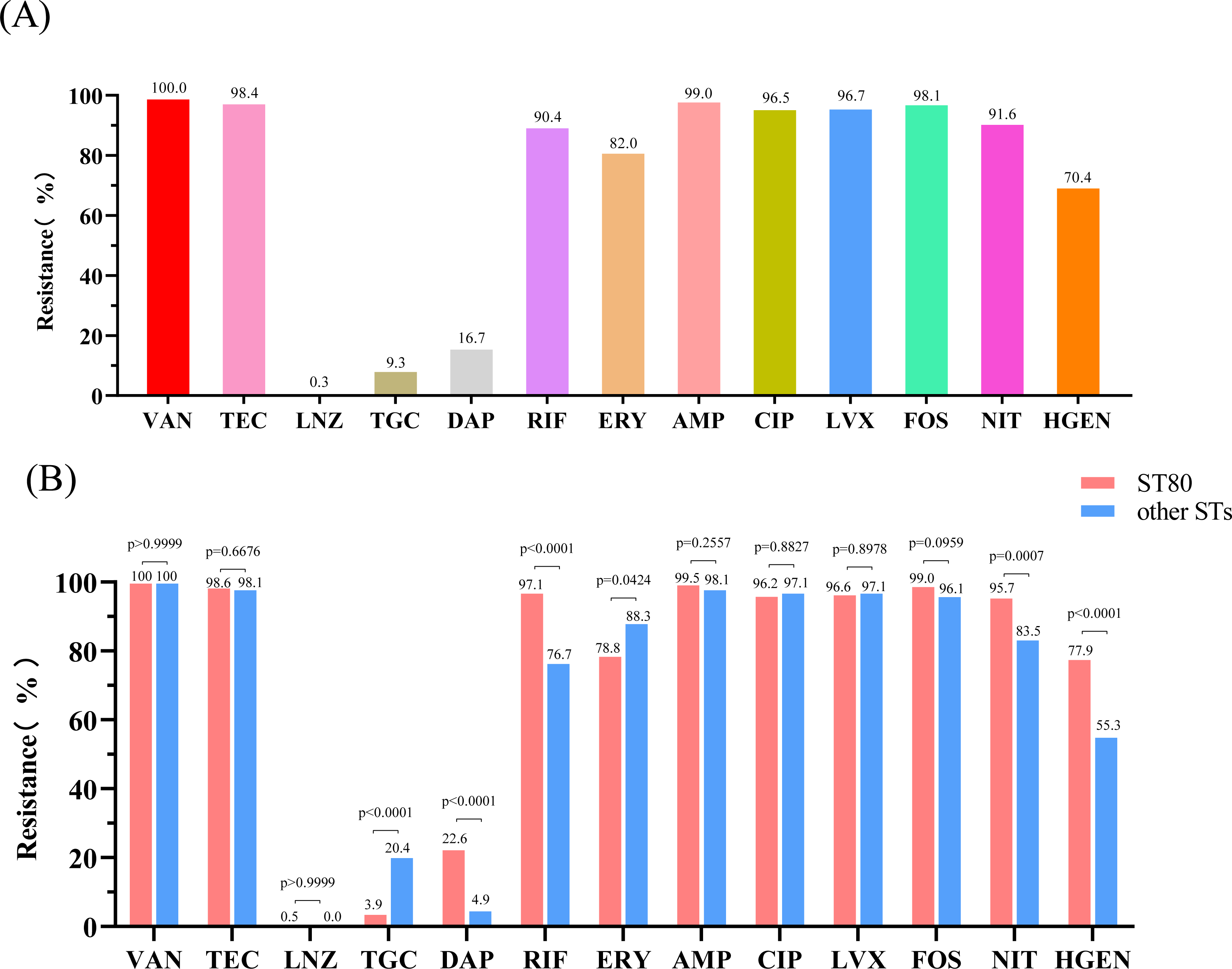
Antimicrobial susceptibility profiles of 13 antimicrobials for 311 VREfm isolates. (A) Overall antimicrobial resistance rate. (B) Antimicrobial resistance rate between ST80 and non-ST80 VREfm. VAN, Vancomycin. TEC, Teicoplanin. LNZ, Linezolid. TGC, Tigecycline. DAP, Daptomycin. RIF, Rifampicin. ERY, Erythromycin. AMP, Ampicillin. CIP, Ciprofloxacin. LVX, Levofloxacin. FOS, Fosfomycin. NIT, Nitrofurantoin. HGEN, High concentration gentamicin.

### 4.4 Population structure and genomic characteristics of VREfm

Pan-genome analysis of 311 VREfm identified 1833 core genes representing a 1.6 Mb alignment in ≥99% of genomes. hierBAPS analysis of VREfm based on 6511 cgSNPs identified 12 sequence clusters (SCs), of which SC12 comprised low-frequency genotypes (Figure 5). ST80 VREfm were distributed into four SCs (SC4, SC5, SC11 and SC12), of which SC11 was the leading lineage. Notably, all ST80 VREfm isolates that were collected after January 2021 belonged to SC11, indicating that the outbreak of VREfm in Guangdong was attributed to SC11. Genetic distance within SC11 showed that they are highly similar (median of cgSNP distance = 6, IQR: 3-8). Six isolates in SC11 were collected from Sichuan province (n=5) and Beijing (n=1) from 2021 to 2023, indicating a potential risk of causing an outbreak beyond Guangdong province.

**Figure 5.**
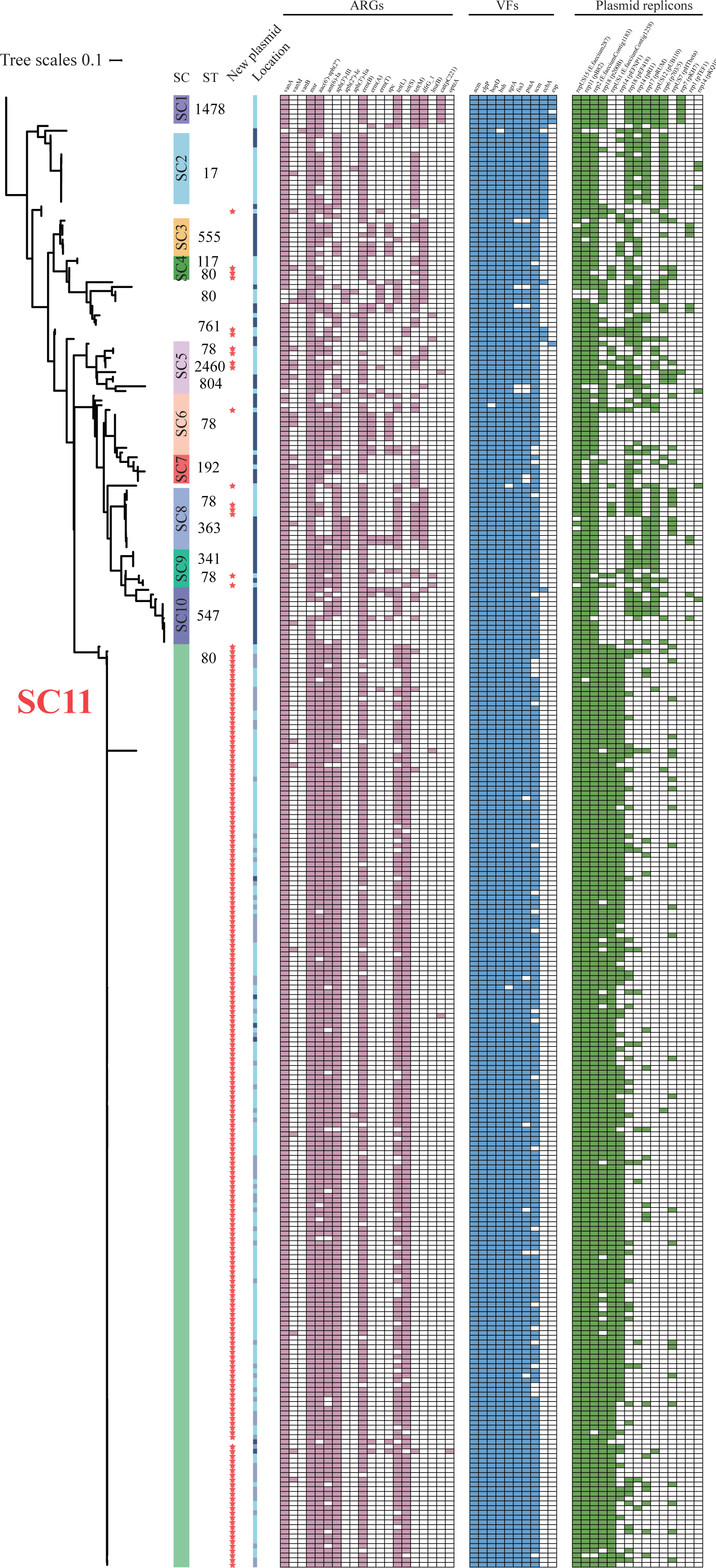
Phylogenetic tree and genomic characteristics of 311 VREfm isolates in this study. The ML phylogeny derived from cgSNPs is at the left of the plot. The lineage colors in the first column denote sequence clusters (SCs). The second column represents the ST. The red star in the third column represents the isolates carried the new type of *vanA*-harboring plasmid. The color strip in the fourth column represents the origin where the isolate from. Light blue represents Guangzhou city; Light purple represents Guangdong province; Dark blue represents other provinces. The heatmaps from left to right represents the presence of antimicrobial resistance genes (pink), virulence factors (blue) and plasmid replicons (green).

A total of 20 ARGs were identified, which attributed to eight antimicrobial classes (Figure 5). 98.07% (306/311) of VREfm isolates harbored *vanA*, of which 7.51% (n=23) co-harbored *vanA* and *vanM*. One isolate only harbored *vanM*. Four isolates harbored *vanB* which was rarely identified in China. Ten VFs and 15 plasmid replicons were verified. However, there is no significant difference in the average number of ARGs, VFs and plasmid replicons between ST80 and other STs VREfm (Supplement figure 3), although the antimicrobial resistance spectrum and MICs varied between ST80 and non-ST80. The distribution of ARGs and plasmid replicons was not significantly different between ST80 and non-ST80 VREfm isolates. Only a virulence gene *ecbA* in the non-ST80 group (17.5%, 18/103) was significantly higher than in the ST80 group (5.8%, 12/208, p=0.0018).

### 4.5 Comparison of ST80 isolates in outbreaks among Guangdong and other countries

To identify the origin and global transmission of ST80 VREfm causing an outbreak in Guangdong province, we additionally included 322 genomes of ST80 VREfm from 22 countries. All the VREfm isolates obtained from Guangdong province exhibit a remarkable convergence within a single lineage (SC11), which was distinctly separated from the isolates found in other countries (Figure 6A). Consistently, this lineage exhibited distinct pan-genome profiles marked by both cgSNPs and accessory genes (Figure 6B). The number of SNPs among ST80 VREfm from China is significantly lower than other countries (p<0.0001), suggesting a clonal transmission (Figure 6C). Notably, one isolate (GCF_012933345.2) collected in 2020 from an Indian patient with a bloodstream infection genetically belonged to this lineage.

**Figure 6.**
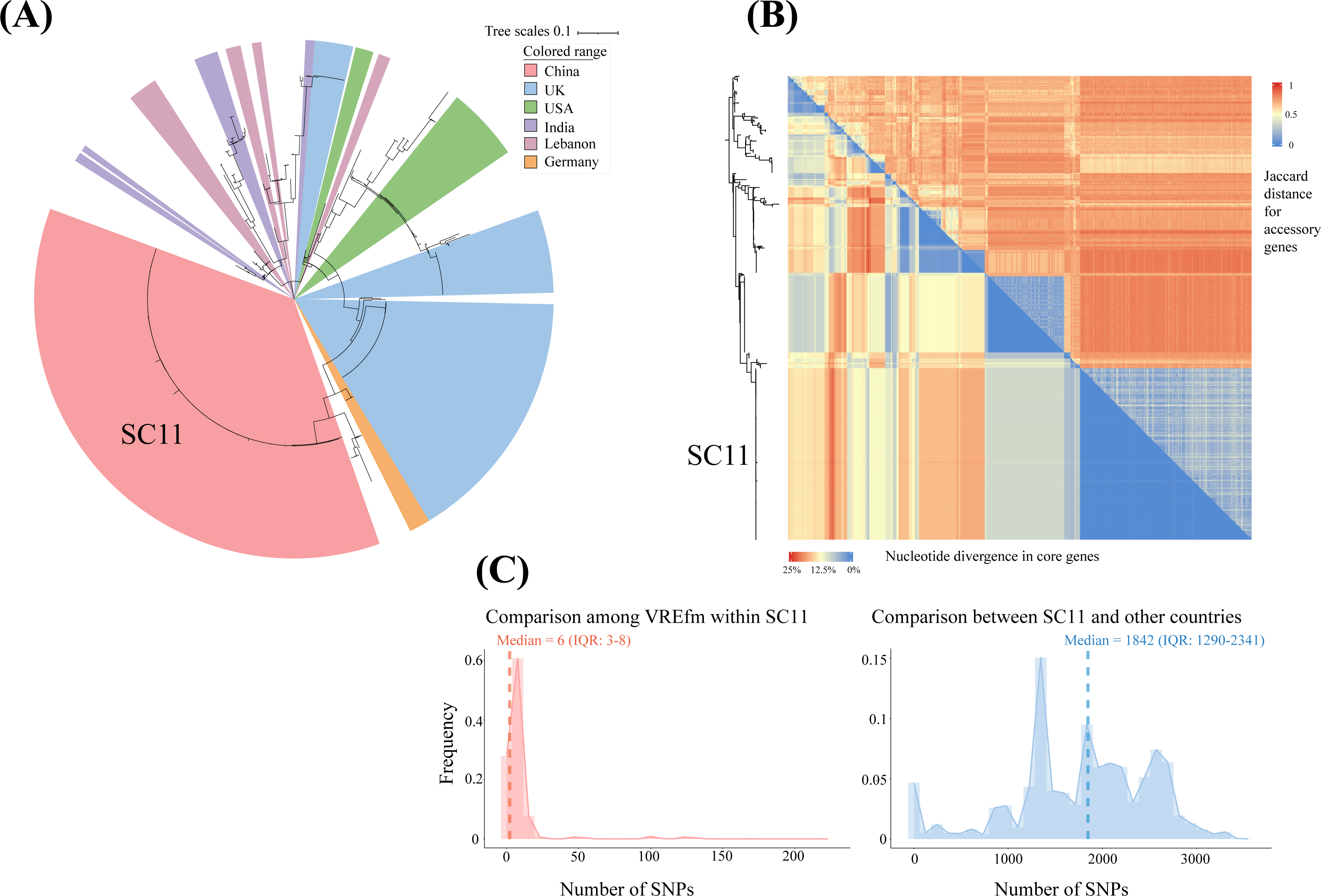
Population structure of ST80 VREfm in different countries. (A) The ML phylogeny was constructed using cgSNPs. The colored branch represents the collected location of the isolate. (B) Matrixes of pairwise distance of core genes (nucleotide divergence, lower left triangle) and accessory genes (Jaccard distance, upper right triangle), ordered against the maximum likelihood phylogeny generated from cgSNPs (left-hand side of panel). The two matrices were highly correlated (p<0.0001, r=0.7727, Mantel test with 1000 permutations). (C) cgSNP distance of ST80 VREfm collected in China and other countries.

Nevertheless, ST80 VREfm are not common in India, indicating that the outbreak of ST80 VREfm in Guangdong may not associated with this strain.

### 4.6 Genetic context and transferability of *vanA*-harboring plasmid in ST80 VREfm

Among 311 VREfm isolates, the *van*-harboring contigs of 100 (32.2%) isolates were matched to the plasmid sequence on GenBank database with >99% identity. However, the *vanA*-harboring contig in 211 (67.8%) of VREfm was not identified against the non-redundant database. One typical isolate (23VRE019) harboring this unique contig was selected for third-generation sequencing, and we noticeably found that this new type of plasmid was identified in all ST80 isolates (n=195) causing outbreaks in Guangdong province from 2021 to 2023 (Figure 5).

Most of the CDSs in p23VRE019 (45,935 bp) were functionally related to IS, plasmid stability and antimicrobial resistance (Figure 7A). This plasmid harbored a *vanA* cassette (*vanRSHAXYZ*) flanked by Tn*1546*/Tn*3* clusters, and a Tn*4001* containing aminoglycosides resistance gene *aac(6’)-aph(2”)*, which was flanked by inverted repeats of IS*256*. No conjugation-related gene was detected in the plasmid. To confirm plasmid transferability, we performed conjugation experiments for all 311 VREfm isolates. Among 100 VREfm carrying *van*-harboring plasmid other than p23VRE019, transconjugants were obtained in 40 isolates. Notably, no transconjugant was obtained among 211 VREfm isolates carrying p23VRE019. Therefore, we speculated that this new *vanA*-harboring plasmid is not conjugative *in vitro* under the experiment condition in this study.

**Figure 7.**
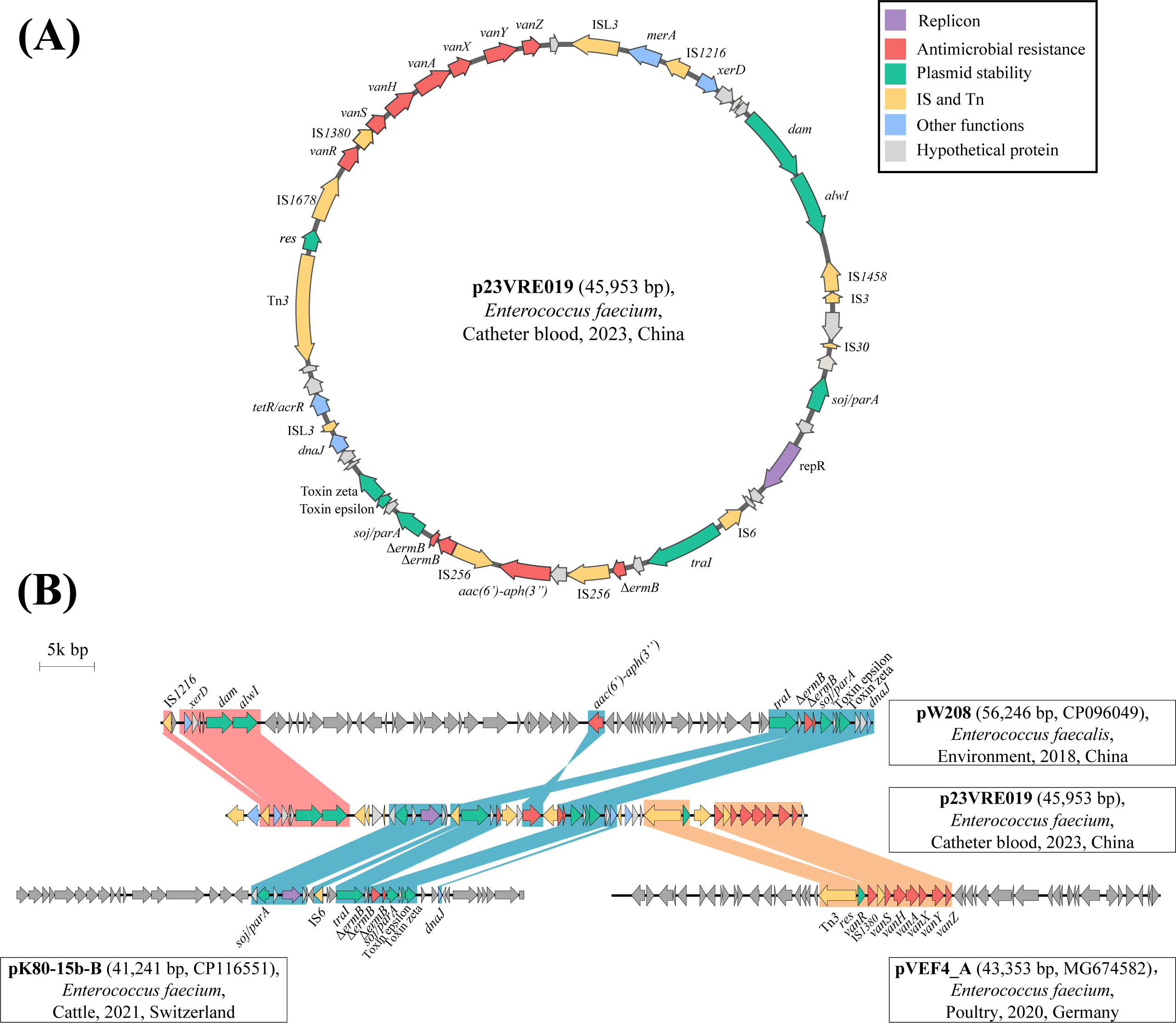
Genomic characteristics of the new type of *vanA*-harboring plasmid. (A) The circle plot of the plasmid profile for p23VRE019. The arrows represent the CDS with its annotated functions. Red, antimicrobial resistance. Green, plasmid stability. Yellow, insertion sequence and transposon. Blue, other functions. Gray, hypothetical proteins. (B) Comparisons of plasmid structures for p23VRE019 and other three plasmids from NCBI. The colored regions represent homologous sequence structures between the plasmids.

Genetic context analysis revealed that p23VRE019 could be formed by three segments from three plasmids (Figure 7B). The plasmid backbone encoded several CDSs associated with plasmid stability and homologised to pW208, which is an environmental *E. faecalis* isolate from China, and pK80-15b-B which is from an *E. faecium* isolate in Switzerland. The *vanA* cassette of p23VRE019 was homologised to pVEF4_A which lack of IS*1678* and IS*L3*. Overall, we concluded that outbreak of *vanA*-positive ST80 VREfm in Guangdong could be attributed to clonal transmission and transposon-mediated horizontal gene transfer of the *vanA* gene cassette.

## 5. Discussion

Emergence and transmission of VREfm pose significant challenges to public health and clinical medicine. Our investigation reveals that the prevalence of VREfm in patients with infection substantially increased in Guangdong province from 2021 to 2023, which was associated with a sub-lineage of ST80 clone harboring a new type of *vanA*-plasmid. To our knowledge, this work represents the largest effort conducted to date to investigate the prevalence and genomic characterizations of VREfm at multicenter scales in China.

In recent decades, the prevalence of VREfm in China has shown low incidence rates, with an average of less than 5%. This is consistent with the findings in other five province, suggesting that the spread of VREfm has not been a significant issue in China [8, 20–23]. However, the dramatic increase of VREfm prevalence in Guangdong province indicated a rapid transmission and outbreak. The monthly epidemiology presented here reflects the pattern with a sharp increase from the beginning of 2021, despite the ongoing COVID-19 containment measures being enforced during the initial half of 2021. Since the disinfectant could facilitate the tolerance of *E. faecium* [31], we speculate that the thrive of VREfm may be associated with the overwhelming use of disinfectants during COVID-19 pandemic. VREfm outbreaks have been reported in healthcare settings, particularly in ICU department, causing various types of healthcare-associated infections including bacteremia [5, 26–29]. It is worth noting that the majority of VREfm isolates were collected from urinary sample from patients suffering from urinary tract infections, indicating that this particular lineage of VREfm could be associated with uropathogenicity.

Several studies have shown that ST17 and ST78 emerged as the predominant types of VREfm in China [8, 22, 24, 25]. Our study, which covers the period from 2014 to 2020, confirmed this observation. However, we noticed a change in the predominant clone of VREfm in Guangdong province since 2021, which is now ST80.

Phylogenetic analysis showed that ST78 and ST80 VREfm in this study are genetically distant, demonstrating that ST80 could be attributed to the introduction of a new clone into hospitals, rather than genome recombination or horizontal gene transfer. ST80 VREfm was identified as a high-risk clone causing VREfm outbreaks in America, Austrillia and Europe countries [5, 26–29]. However, the ST80 VREfm isolates in our study (SC11) were genetically distinct from ST80 VREfm found in other countries. The only identical ST80 VREfm in India was identified in the Genbank database, but it is not the predominant type in India. These results implied that this sub-lineage could be sporadically distributed around the world, but certain factors could trigger and facilitate its transmission, causing outbreaks in Guangdong, China. The pan-genome analysis conducted on the isolates of SC11 signified a high degree of genomic diversity, plasticity and unique attributes, implying a potential to spreading and becoming a prevalent clone.

Based on the results of multiple antimicrobial susceptibility tests, the ST80 VREfm exhibits to have a significantly high level of resistance to vancomycin compare to non-ST80 isolates. Currently, daptomycin and linezolid are the most utilised “last- line” antibiotics for the treatment of VRE infections [1]. However, our research reveals that a significant percentage of ST80 VREfm is resistant to daptomycin, which limits treatment options. Conversely, only one single isolate was found to be resistant to linezolid, demonstrating its effectiveness against ST80 VREfm.

The vancomycin resistance mechanisms of ST80 VREfm isolates caused outbreak in other countries mostly attributed to *vanA* or *vanB* [5, 26, 27]. In this study, all VREfm isolates in Guangdong harbored a *vanA* gene flanked by Tn*1546*, which is consistently the predominant mechanism of VREfm in China [8]. *vanM* gene was first found in VREfm in Shanghai in 2006 and is locally prevalent in China [30]. In this study, a small proportion of *vanA*-harboring ST80 VREfm co-carried *vanM*, suggesting a rapid genomic interaction occurring in various lineages of ST80 VREfm.

We found a new type of *vanA*-harboring plasmid associated with the outbreak clone of ST80-type VREfm in Guangdong. Notably, this plasmid also detected in various STs such as ST78, ST17, implying that there is an interaction and transmission of this *vanA*-plasmid among VREfm populations. However, the sequence analysis and plasmid conjugation experiments showed that this plasmid is not conjugative, at least *in vitro* and under the conditions tested in this study. This means that the dissemination and outbreak of *vanA*-harboring ST80 VREfm is likely caused by acquisition of insertion sequence element and clonal transmission. This new type of plasmid has been detected in ST80 VREfm from Beijing and Sichuan province, indicating a potential risk of spreading and outbreak in the future.

Our study has several limitations. First, while our study has shed light on the emergence and dramatic increase of ST80 VREfm in Guangdong, the genomic epidemiological data we have presented may not be generalisable throughout China. According to data published by CHINET, the prevalence of VREfm increased solely in Guangdong province from January to June in 2023, which is in line with the observations made in our study. Second, the underlying contributors to the rapid transmission and outbreak of ST80 VREfm have yet to be determined, especially the measures to contain COVID-19 were still being enforced during the initial half of 2021. Further case-control studies, risk factor analyses and metagenomic investigation would be helpful to bridge this gap. Finally, the pathogenicity of ST80 VREfm, and the stability and fitness cost of this new plasmid were not resolved, and are part of future work.

Despite of these limitations, our study revealed an ongoing outbreak of ST80 VREfm with a new type of *vanA*-harboring plasmid in Guangdong province. It is concerning that this outbreak clone has also been detected in several provinces in China and other countries, foreboding a potential risk that the prevalence of VREfm may increase in the coming years. Continuous surveillance is needed to monitor the prevalence, transmission, and outbreak of this high-risk ST80 VREfm clone to inform public health officials and adapt infection prevention and control interventions.

## Author contributions

CS, LL, and YX contributed equally in this study. CS drafted the first version of the manuscript, which was reviewed and edited by CC and BH. CS, LL, and YX were responsible for concept, bioinformatical and statistical analyses. CS, LL, YX, HZ, JZ, LZ, JP, and JZ were responsible for the data collection and did the experiments. CS, LL, YX, CC, BH, NZ, YJ, DC, GL, KW, MW, XG and JW were responsible for the sample collection. All authors had full access to all the data in the study and took responsibility for the integrity of the data and the accuracy of the data analysis. All authors reviewed, revised, and approved the final submission.

## Transparency declaration

The authors declare no conflict of interest.

## Data Availability

The genome assemblies of VREfm reported in this study have been deposited in the NCBI GenBank genomic DNA database under BioProject accession number PRJNA1003636.

## Acknowledgements

This work was supported by the National Natural Science Foundation of China (grant numbers 82302598 to CS), Guangzhou Basic and Applied Foundation (grant number 2023A04J0456 to CS), Guangdong Basic and Applied Research Foundation (grant number 2022A1515111171 to CS), China Postdoctoral Science Foundation (grant numbers 2023T160150 and 2022M720922 to CS), Guangdong Provincial Hospital of Chinese Medicine (grant numbers YN2022QN11 to CS), Guangdong Provincial Key Laboratory of Research on Emergency in TCM (2023B1212060062 to CS).

## Supplementary figure legends

Supplementary figure 1. Distribution of location (A) and source (B) of 311 VREfm isolates in minimum-spanning tree.

Supplementary figure 2. Comparisons of MICs for 13 antimicrobials between ST80 and non-ST80 isolates.

Vertical dash lines indicate the breakpoint of resistance according CLSI. Drug abbreviations are as per figure 4.

Supplementary figure 3. Comparisons of ARGs, VFs and plasmid replicons between ST80 and non-ST80 VREfm.

The calculations of statistical difference were done using student *t* test.

