## Supplement figure 1 for "Emergence and ongoing outbreak of ST80 vancomycin-resistant *Enterococcus faecium* in Guangdong province, China from 2021 to 2023: a multicenter, time-series and genomic epidemiological study"

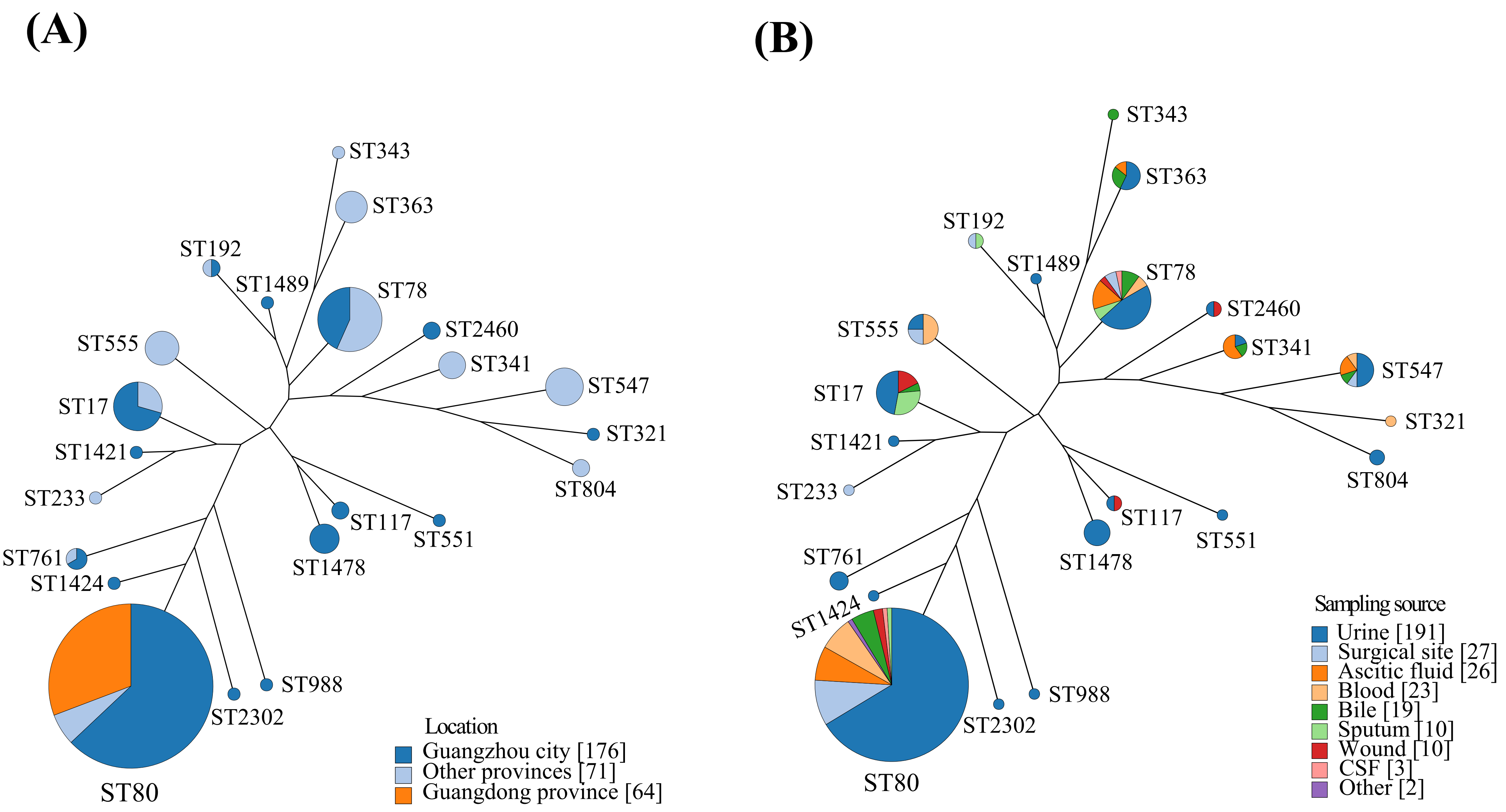


**Supplementary figure 1. Distribution of location (A) and source (B) of 311 VREfm isolates in minimum-spanning tree.**
