## Supplementary figures and images for "Emergence and ongoing outbreak of ST80 vancomycin-resistant *Enterococcus faecium* in Guangdong province, China from 2021 to 2023: a multicenter, time-series and genomic epidemiological study"

### Supplement figure 2

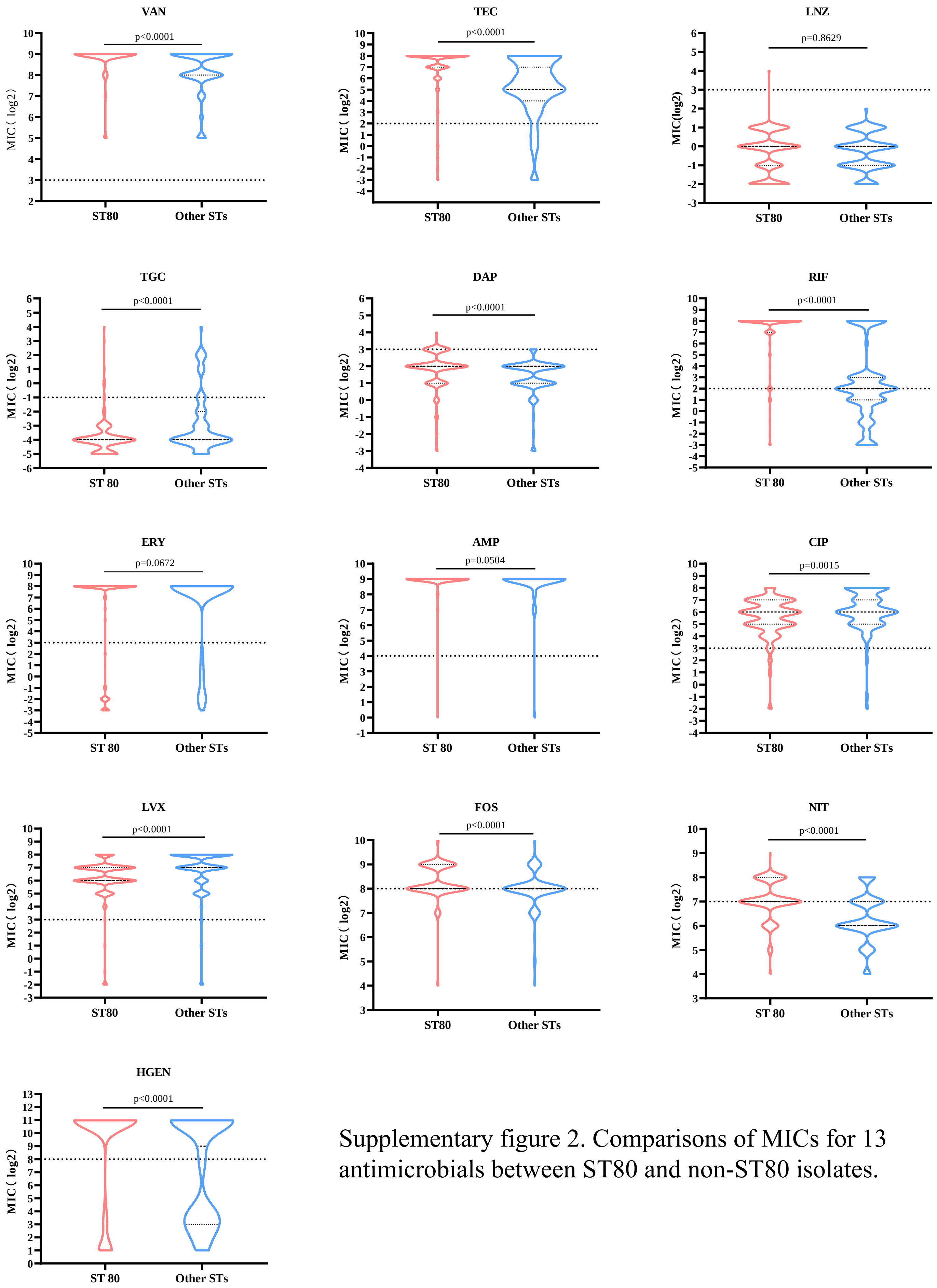


Supplementary figure 2. Comparisons of MICs for 13 antimicrobials between ST80 and non-ST80 isolates.
