## Supplement figure 3 for "Emergence and ongoing outbreak of ST80 vancomycin-resistant *Enterococcus faecium* in Guangdong province, China from 2021 to 2023: a multicenter, time-series and genomic epidemiological study"

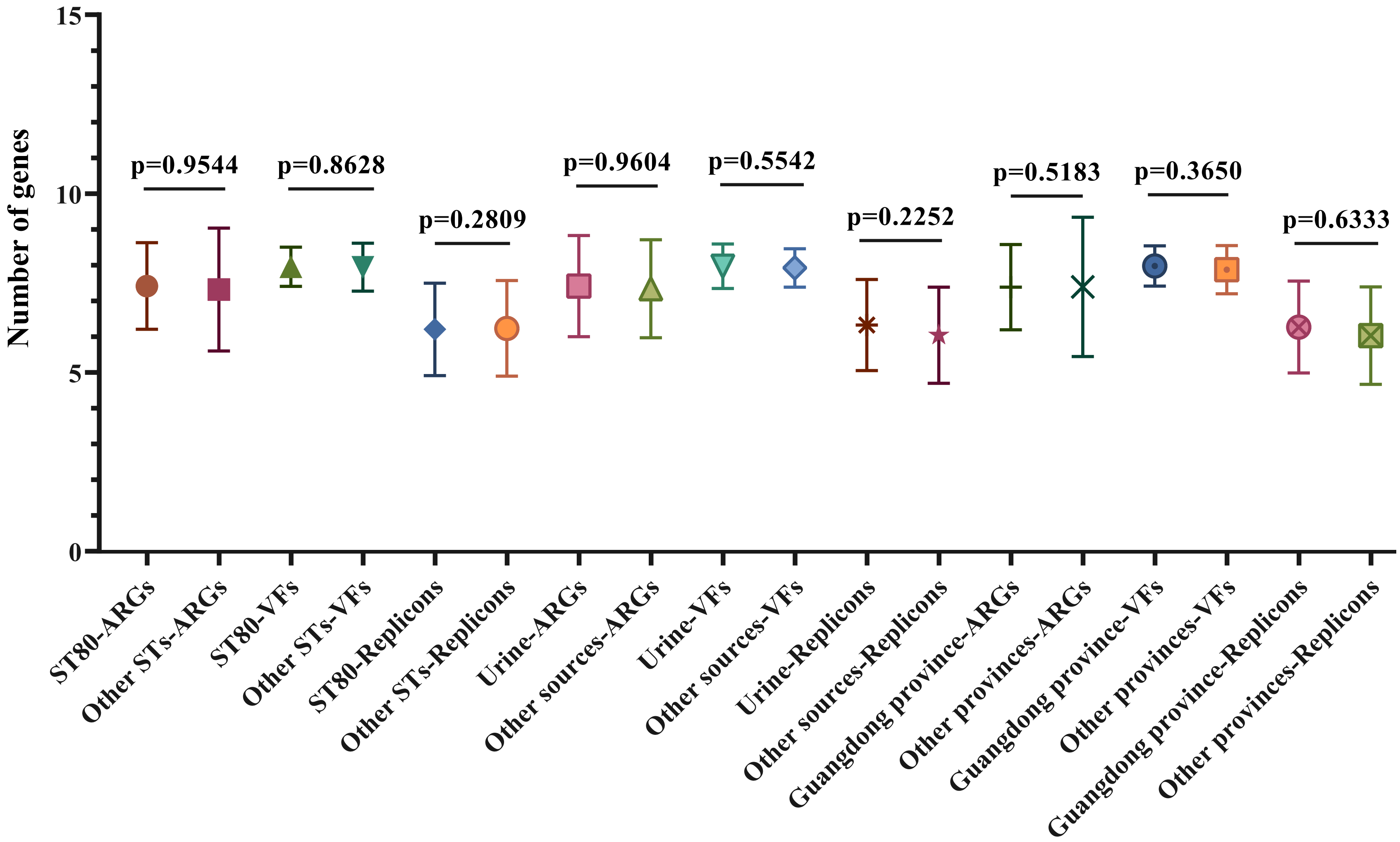


**Supplementary figure 3. Comparisons of ARGs, VFs and plasmid replicons between ST80 and non-ST80 VREfm.**
